# Patterns of emergency department use among young people with bipolar disorder: A data linkage cohort study

**DOI:** 10.64898/2026.05.07.26352617

**Authors:** Ashlee Turner, Ian B Hickie, Mathew Varidel, Nicholas Ho, Catherine M McHugh, Jacob J Crouse, Joanne S Carpenter, Alissa Nichles, Natalia Zmicerevska, Yun Ju (Christine) Song, Elizabeth M Scott, Frank Iorfino

## Abstract

**Objective:** To charactertise emergency department (ED) use among young people with bipolar disorder (BD) and compare patterns to those observed in anxiety, depressive, and psychotic disorders.

**Design, setting and participants:** Data linkage study using administrative ED presentation records (January 2020 to October 2020) and a transdiagnostic youth mental health cohort of 2243 individuals aged 12–30 years in New South Wales, Australia.

**Main outcome measures:** ED presentation patterns (any presentation, frequency, and rates) and reasons for presentation (mental health-related and non-mental health-related).

**Results:** Of the 354 young people with BD, 309 (87.3%) presented to an ED at least once. ED presentation rates were higher for BD than for anxiety (incidence rate ratio [IRR]=1.82, p<.001) and depressive disorders (IRR=1.32, p<.001), but similar to psychotic disorders (IRR=0.91, p=.379). Differences were primarily driven by mental health-related presentations. Recurrent mental health presentations were associated with illness progression (clinical stage and functional impairment) rather than diagnosis. However, the likelihood of mental health-related presentations remained higher in BD compared with anxiety and depressive disorders after adjustment.

**Conclusions:** Young people with BD have high rates of ED use, comparable to those with psychotic disorders. Although mental health-related presentations are more common in BD than in anxiety and depressive disorders, recurrence is largely explained by markers of illness progression. These findings highlight the need for community-based services that provide continuous and coordinated care for young people with complex mental health needs.

## 1. INTRODUCTION

Bipolar disorder (BD), a chronic, severe mental disorder characterised by recurrent depressive and (hypo)manic episodes, typically emerges during adolescence and early adulthood. It is associated with substantial functional and cognitive impairment, even in early illness stages [1]. Earlier onset predicts a more severe clinical course, poorer long-term outcomes, and higher rates of suicidal behaviour [2, 3]. Among individuals aged 10-24, BD is the fourth leading cause of disability worldwide [4], highlighting its contribution to disease burden during this critical developmental period.

Despite frequent health service use [5], many people receive suboptimal care [6] and subsequent presentations to emergency departments (EDs) may signal periods of acute escalation and gaps in continuity of care. Approximately three in four young people presenting to EDs with BD have prior mental health service use [7], indicating that ED use occurs despite service engagement. While EDs are critical for managing acute mental health crises, they cannot provide the coordinated, continuous care needed for chronic conditions like BD.

Reported ED presentation rates among young people with BD vary widely (10-20%) [8, 9], and many studies rely on insurance claims data. These data are limited by diagnostic uncertainty in youth [10] and restricted generalisability to insure populations, potentially excluding marginalised groups [11]. Studies using electronic medical records of clinically diagnosed cohorts address some of these issues. For example, nearly half of 85 young people (aged 6-17) presented to a psychiatric ED within two years of diagnosis, averaging almost two presentation per year despite complex medication regimes [12]. Another study of 100 young people (aged 12-18) reported 19% presenting to a psychiatric ED and 39% to a general ED [5]. While these approaches overcome the limitations of previous studies and provide valuable clinical insight, they are limited by small sample sizes and lack comparison with other diagnostic groups commonly seen in youth.

Comparative analyses are needed to determine whether ED use patterns in BD reflect diagnosis-specific needs or broader service use trends. Anxiety and depressive disorders are highly prevalent among help-seeking youth [13] and are associated with high ED use [14], and psychotic disorders are typically associated with increased acute service use [15]. Comparing ED use across these groups is essential to clarify relative healthcare needs and inform planning.

Youth mental health service cohorts offer a unique opportunity to address these gaps by providing access to diverse, help-seeking populations. Prior work (based on the same cohort as the present study) shows that three quarters of young people engaged in such services have also used EDs for mental or physical health problems [16]. However, ED presentation patterns specific to BD across all presentation types, and their comparison with other diagnostic groups, remain unexplored. Building on these previous findings, this study aimed to: (1) characterise ED presentation patterns among young people with BD within a youth mental health cohort with linked service utilisation data; and (2) compare these patterns across anxiety, depressive, and psychotic disorders using a large linked dataset of ED presentations in New South Wales (NSW), Australia.

## 2. METHODS

### 2.1 Participants and setting

Participants were drawn from the Brain and Mind Centre’s Patient Research Register (BPRR), a research register of 7024 individuals aged 12-30 years old who presented to early intervention youth mental health clinics in Sydney (Australia) between October 2008 and October 2020. Exclusion criteria were: (1) medical instability or inability to give informed consent; (2) history of neurological disease (e.g., epilepsy); (3) medical illness known to affect cognitive and brain function (e.g., cancer, electroconvulsive therapy in the previous 3 months); (4) clinically evident intellectual disability; and/or (5) insufficient English proficiency.

A subset of 2868 individuals were tracked longitudinally as they accessed care in the service. For the purposes of the current analysis, data were taken from each participant’s most recent clinical assessment and included individuals with a primary diagnosis of BD, anxiety, depressive, or psychotic disorder. This approach was chosen to capture the most current and clinically relevant information. A flow diagram of inclusion and exclusion is included in the Supplementary Material (Figure S1).

### 2.2 Data collection

A detailed description of the clinical data collection process is described elsewhere [13]. Briefly, demographic, clinical, and functional data were retrospectively extracted from clinical files using a standardised clinical proforma. For each participant, the first clinical assessment at the service was defined as the baseline timepoint, and its date determined each follow-up time points (set at 3 months, 6 months, 12 months, 2 years, 3, years, 4 years, and 5 years post-baseline). For this study, relevant measures included: demographic characteristics (age, sex); mental disorder diagnoses classified according to the Diagnostic and Statistical Manual of Mental Disorders (DSM-5) criteria [17]; clinical stage assessed according to a previously established model and assigned as Stage 1a (help-seeking participants with mild to moderate, non-specific symptoms), Stage 1b (attenuated syndromes with specific symptoms that may meet diagnostic criteria for specific disorders), or Stage 2+ (discrete, full-threshold disorders with moderate-severe persistent symptoms); and social and occupational functioning assessed with the Social and Occupational Functioning Assessment Scale (SOFAS) [18].

### 2.3 Data linkage

The BPRR was linked to three external databases held by the Centre for Health Record Linkage in New South Wales (NSW), Australia. The complete details of the data linkage approach have been published elsewhere [19]. Clinical research data for this cohort were linked to routinely collected data from the ED Data Collection (EDDC) registry over a 10-year period, from January 2010 to October 2020.

This study reports on data from the EDDC registry, which contains routinely collected data for presentations to 184 public hospital EDs in NSW. Reasons for presentation were classified into predetermined categories by two clinically trained authors using SNOMED, ICD-9 and ICD-10 codes: (a) mental illness; (b) suicidal behaviours and self-harm; (c) alcohol and substance misuse; (d) accident and injury; (e) physical illness; and (f) other. Interrater reliability was 98% with all discrepancies between the coders resolved via consensus discussion.

### 2.4 Statistical analysis

Analyses were conducted in R (version 4.4.1) [21] . Descriptive data are presented as mean ± standard deviation (SD) or median (interquartile range [IQR]) for continuous variables and frequencies (%) for categorical data.

The frequency and rate of ED presentations were analysed in a stepwise manner to compare utilisation patterns across diagnostic groups. Chi-square tests of independence were used to examine overall associations between mental health diagnosis and: (a) any ED presentation (yes/no), and (b) presentation frequency patterns (single versus multiple presentations). Post-hoc analyses were conducted using pairwise chi-square tests for significant differences, and odds ratios with 95% confidence intervals (95% CIs) were calculated to quantify the magnitude of associations. Negative binomial regression (to account for overdisperson) (MASS package [version 7.3.65] [22]) was used to model ED presentation rates across diagnostic groups. Incidence rate ratios (IRRs) with 95% CIs were calculated using BD as the reference category.

Logistic regression models (base R glm() function) were used to examine associations between diagnostic groups and reasons for ED presentation. Odds ratios with 95% CIs were calculated to quantify the magnitude of association between diagnostic group and presentation reasons. Two primary outcome variables were created by combining related presentation reasons: (1) any mental health-related presentation (mental illness, alcohol or substance misuse, or suicidal behaviours or self-harm); and (2) any non-mental health presentation (accident or injury or physical illness).

All analyses were conducted in two stages, with BD as the reference diagnostic category: (1) unadjusted models; and (2) adjusted models controlling for age at first ED presentation, sex, clinical stage, and level of functioning. For interpretability, all IRRs and ORs were inverted (1 / x), such that values >1 indicate higher odds/rates for the BD group compared to other diagnostic groups.

### 2.5 Ethics approval

The BPRR was approved by the University of Sydney’s Human Research Ethics Committee (2008/5453, 2012/1626). The data linkage study was approved by the NSW Population and Health Services Research Committee (2019/ETH12201).This included approval of a waiver of consent according to the National Statement on Ethical Conduct in Human Research (2007) [20].

## 3. RESULTS

A total of 2243 participants met eligibility criteria and were included in the final analysis (61.35% female; mean age ± SD = 21.32 ± 4.16). Table 1 presents demographic and clinical characteristics at the most recent service presentation.

**Table 1.** Demographic and clinical characteristics of the sample at last mental health service presentation.

| Characteristic | Total sample (n=2243) | Anxiety disorder (n=601) | Bipolar disorder (n=354) | Depressive disorder (n=1066) | Psychotic disorder (n=222) |
| --- | --- | --- | --- | --- | --- |
| Age, years | $21.32 \pm 4.16$ | $20.61 \pm 3.80$ | $22.70 \pm 4.08$ | $20.74 \pm 3.95$ | $23.87 \pm 4.69$ |
| Sex, % female | 1376 (61.35) | 364 (60.57) | 251 (70.90) | 692 (64.92) | 69 (31.08) |
| Clinical stage<sup>a</sup> |  |  |  |  |  |
| Stage 1a | 352 (15.69) | 181 (30.12) | 6 (1.69) | 165 (15.48) | 0 (0.00) |
| Stage 1b | 1384 (61.70) | 398 (66.22) | 165 (46.61) | 771 (72.33) | 50 (22.52) |
| Stage 2+ | 482 (21.49) | 21 (3.49) | 175 (49.44) | 122 (11.44) | 164 (73.87) |
| SOFAS, score /100 | 63.15 ± 11.00 | 65.96 ± 9.46 | 61.51 ± 12.52 | 63.81 ± 10.21 | 54.95 ± 11.68 |

Data are presented as mean ± standard deviation for continuous variables and n (%) for categorical variables.
Abbreviations: SOFAS = Social and Occupational Functioning Assessment Scale.
<sup>^</sup>n=25 excluded due to 'uncertain' or missing clinical stage.

### 3.1 Rate and frequency of ED presentations

Of the 2243 individuals, 1800 (80.25%) had at least one ED presentation during the study period (Table 2). The demographic and clinical characteristics of those who presented are summarised in Table S1. Diagnostic groups differed in the likelihood of ED presentation (χ²=45.89, df=3, p<.001) (Figure 1). Pairwise comparisons showed that young people with BD were more likely to present (median presentations: 5) than those with anxiety (median presentations: 3, OR=2.66, 95% CI [1.86, 3.82], p<.001) and depressive disorders (median presentations: 4, OR=1.63, 95% CI [1.15, 2.30], p=.045). There was no difference in the likelihood of presentation between those with BD and psychotic disorders (median presentations: 5, OR=0.91, 95% CI [0.54, 1.52], p=1.00).

**Figure 1.**
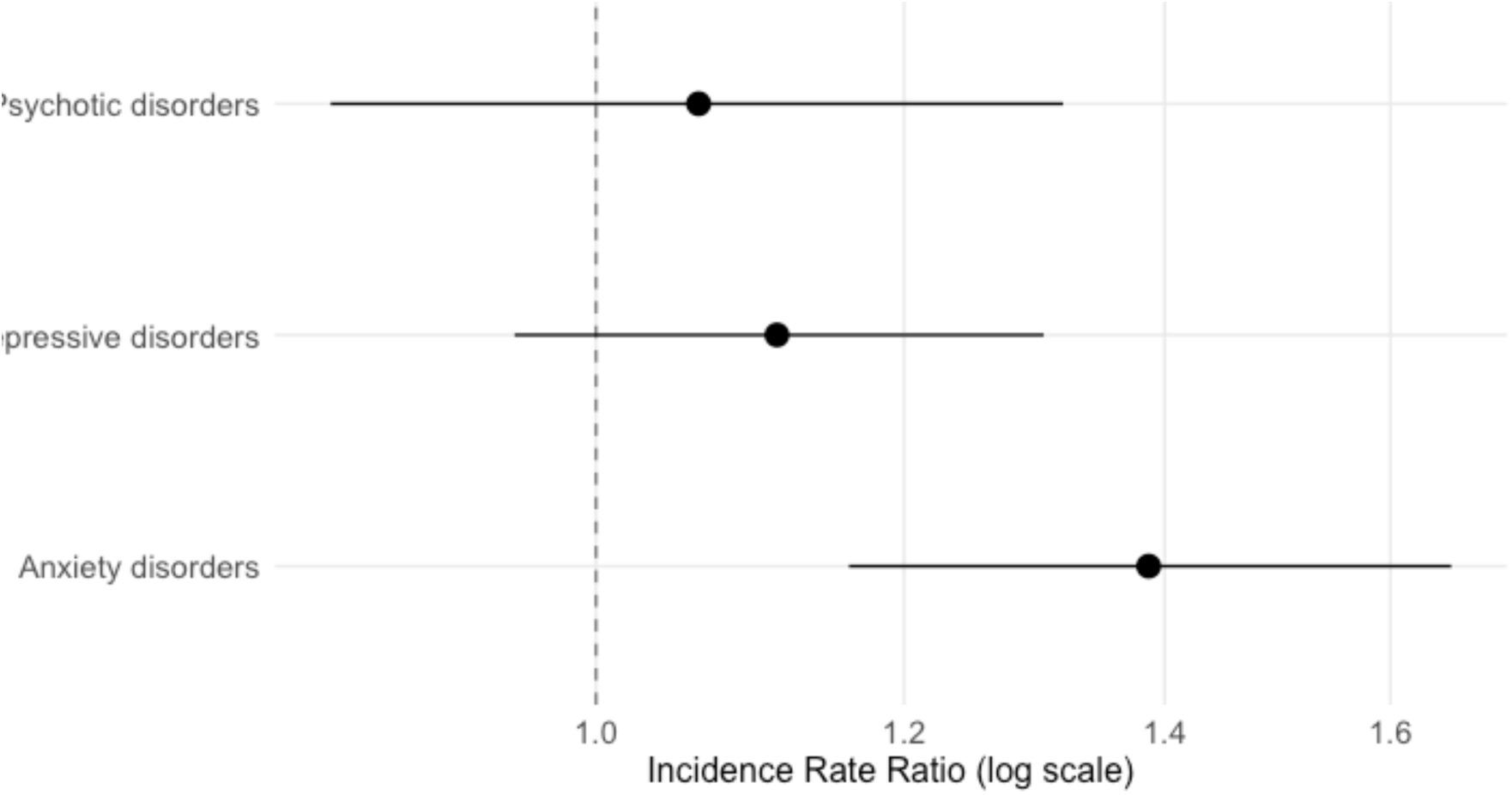
Adjusted incidence rate ratios for ED presentations by diagnostic group. Note: bipolar disorder is the reference category; error bars represent 95% confidence intervals, n=2218 (n=25 excluded due to ‘uncertain’ or missing clinical stage).

**Table 2.** Incidence rate ratios for ED presentation of those with bipolar disorder compared with other diagnostic groups (complete sample)

| Diagnostic group | Unadjusted IRR [95% CI] | Adjusted IRR [95% CI]<sup>a</sup> |
| --- | --- | --- |
| Anxiety disorder | 1.82 [1.54, 2.15]** | 1.39 [1.16, 1.66]** |
| Depressive disorder | 1.32 [1.13, 1.53]** | 1.11 [0.95, 1.31] |
| Psychotic disorder | 0.91 [0.73, 1.12] | 1.06 [0.86, 1.32] |

<sup>a</sup>Adjusted for sex, clinical stage, and functioning.
Abbreviations: IRR = Incidence Rate Ratio; CI = Confidence Interval.
n=2218 (n=25 excluded due to 'uncertain' or missing clinical stage).
Reference group: Bipolar disorder.
\* $p<.05$ , \*\* $p<.001$

**Figure 2.**
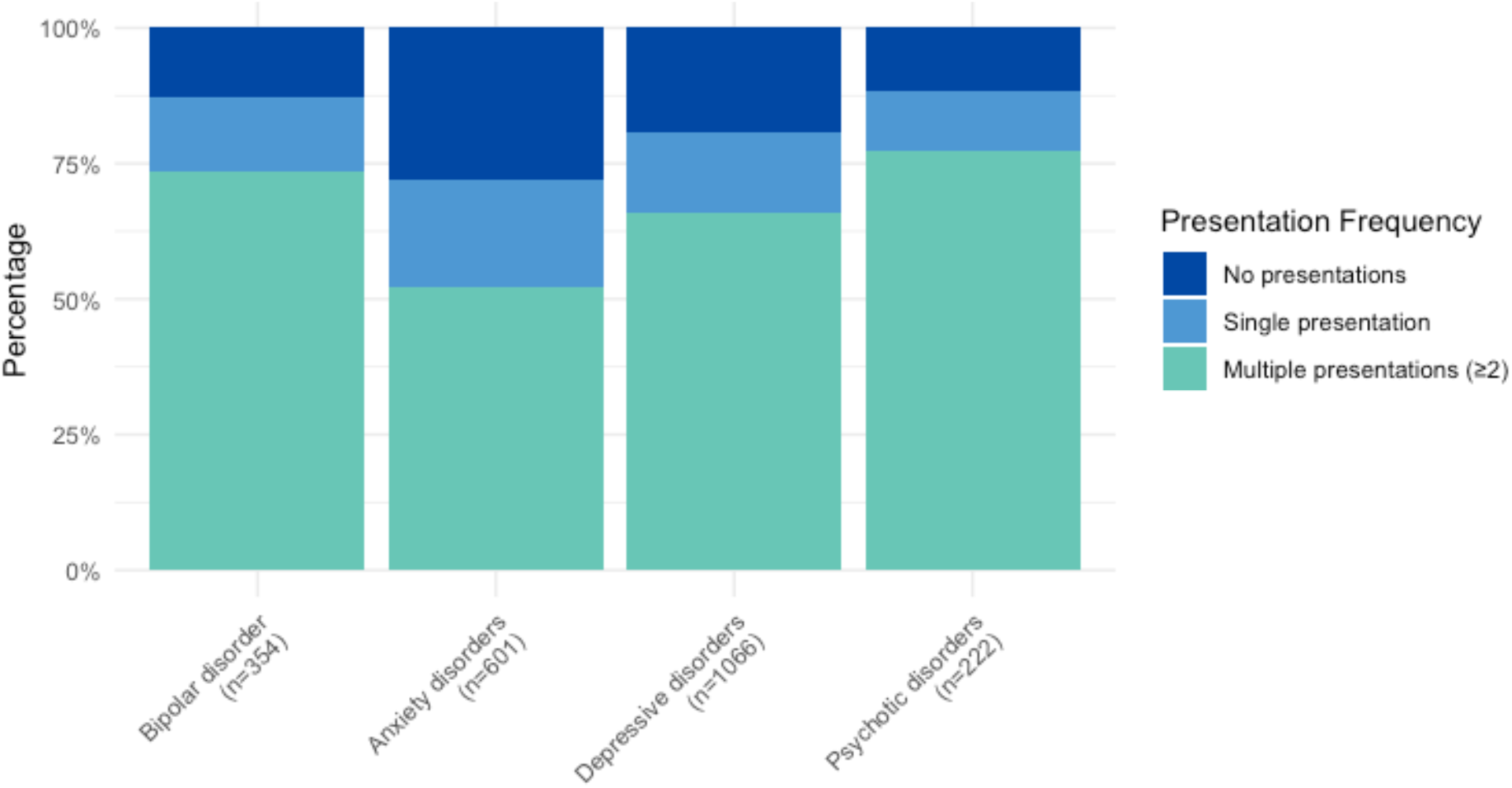
Distribution of ED presentation frequency across diagnostic groups.

Unadjusted negative binomial regression showed higher ED presentation rates in young people with BD compared with anxiety (IRR=1.82, 95% CI [1.54, 2.15], p<.001) and depressive disorders (IRR=1.32, 95% CI [1.13, 1.53], p<.001) (Table 3), with no difference versus psychotic disorders (IRR=0.91, 95% CI [0.73, 1.12], p=.372). After adjusting for sex, clinical stage, and functioning, the association remained significant only for anxiety disorders (IRR=1.39, 95% CI [1.16, 1.66] p<.001), with no differences for depressive (IRR=1.11, 95% CI [0.95, 1.31], p=.188) or psychotic disorders (IRR=1.06, 95% CI[0.86, 1.32], p=.581) relative to BD (Figure 1). Female sex, greater functional impairment, and more advanced clinical stage (Stage 2+) were independently associated with higher ED presentation rates (Table S2).

**Table 3.** Distribution of ED presentations by diagnostic group.

| Diagnostic group | Any ED presentation | Single presentation | Multiple presentations ($\geq 2$) | Total presentations |
| --- | --- | --- | --- | --- |
| Bipolar disorder | 309 (87.29) | 49 (15.86) | 260 (84.14) | 5 (2.0 – 10.0) |
| Anxiety disorder | 433 (72.05) | 119 (27.48) | 314 (72.52) | 3 (1.0 – 6.0) |
| Depressive disorder | 862 (80.86) | 158 (18.33) | 704 (81.67) | 4 (2.0 – 8.0) |
| Psychotic disorder | 196 (88.29) | 24 (12.24) | 172 (87.76) | 5 (2.0 – 10.0) |
| Total | 1800 (80.25) | 350 (19.44) | 1450 (80.56) | 4 (2.0 – 8.0) |

Data are presented as n (%) for categorical variables and median (IQR) for continuous variables.

Among those with ≥1 ED presentation, diagnostic groups differed in the likelihood of single versus multiple presentations (>2) (χ²=27.57, df=3, p<.001). Young people with BD were twice as likely to present multiple occasions, compared to those with anxiety disorder (OR=2.01, 95% CI [1.39, 2.93], p=.002). No differences were observed between BD and depressive (OR=1.19, 95% CI [0.84, 1.70], p=1.00) or psychotic disorders (OR=0.74, 95% CI [0.43, 1.25], p=1.00).

### 3.2 Reasons for presentation

Of the 309 young people with BD who presented to an ED, 190 (61.49%) presented for a mental health-related reason at least once (Table S3). Individuals with BD accounted for 23.33% of mental health-related presentations, despite representing 17.17% of ED users. They had higher odds of mental health-related presentations than those with anxiety and depressive disorders in both unadjusted and adjusted models, with no difference versus psychotic disorders (Table 4).

**Table 4.**
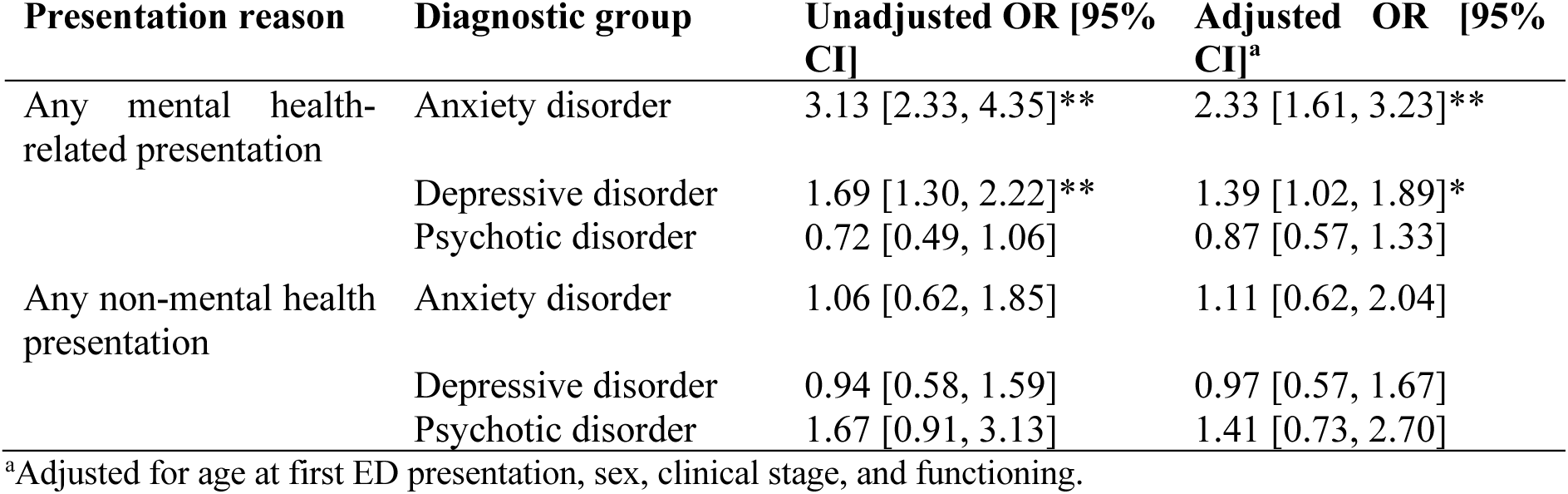
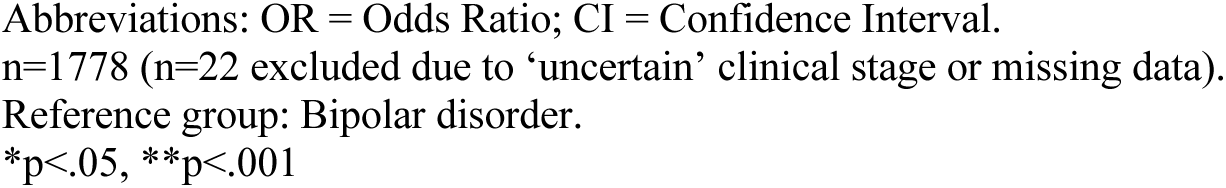
Composite reasons for ED presenations those with bipolar disorder compared with other diagnostic groups (anxiety, depressive, psychotic).

| Presentation reason | Diagnostic group | Unadjusted OR [95% CI] | Adjusted OR [95% CI]<sup>a</sup> |
| --- | --- | --- | --- |
| Any mental health-related presentation | Anxiety disorder | 3.13 [2.33, 4.35]** | 2.33 [1.61, 3.23]** |
|  | Depressive disorder | 1.69 [1.30, 2.22]** | 1.39 [1.02, 1.89]* |
|  | Psychotic disorder | 0.72 [0.49, 1.06] | 0.87 [0.57, 1.33] |
| Any non-mental health presentation | Anxiety disorder | 1.06 [0.62, 1.85] | 1.11 [0.62, 2.04] |
|  | Depressive disorder | 0.94 [0.58, 1.59] | 0.97 [0.57, 1.67] |
|  | Psychotic disorder | 1.67 [0.91, 3.13] | 1.41 [0.73, 2.70] |

<sup>a</sup>Adjusted for age at first ED presentation, sex, clinical stage, and functioning.
Abbreviations: OR = Odds Ratio; CI = Confidence Interval. n=1778 (n=22 excluded due to ‘uncertain’ clinical stage or missing data). Reference group: Bipolar disorder. \*p<.05, \*\*p<.001

In adjusted analyses, younger age at first ED presentation, greater functional impairment, and more advanced clinical stage were independently associated with higher odds of mental health-related presentations (Table S4). Diagnostic groups did not differ in non-mental health presentations in unadjusted or adjusted analyses (all p>.05, Table S5).

Of the 190 young people with BD who presented a for mental health-related reason, 125 did so multiple times (65.79%; Figure 3). Unadjusted analyses showed higher odds of multiple mental health-related presentations in BD compared with anxiety (OR=1.71, 95% CI [1.10, 2.68], p=.018) and depressive disorders (OR=1.62, 95% CI [1.13, 2.33], p=.009), with no difference versus psychotic disorders (OR=0.77, 95% CI [0.47, 1.25], p=.294) (Table 5).

**Figure 3.**
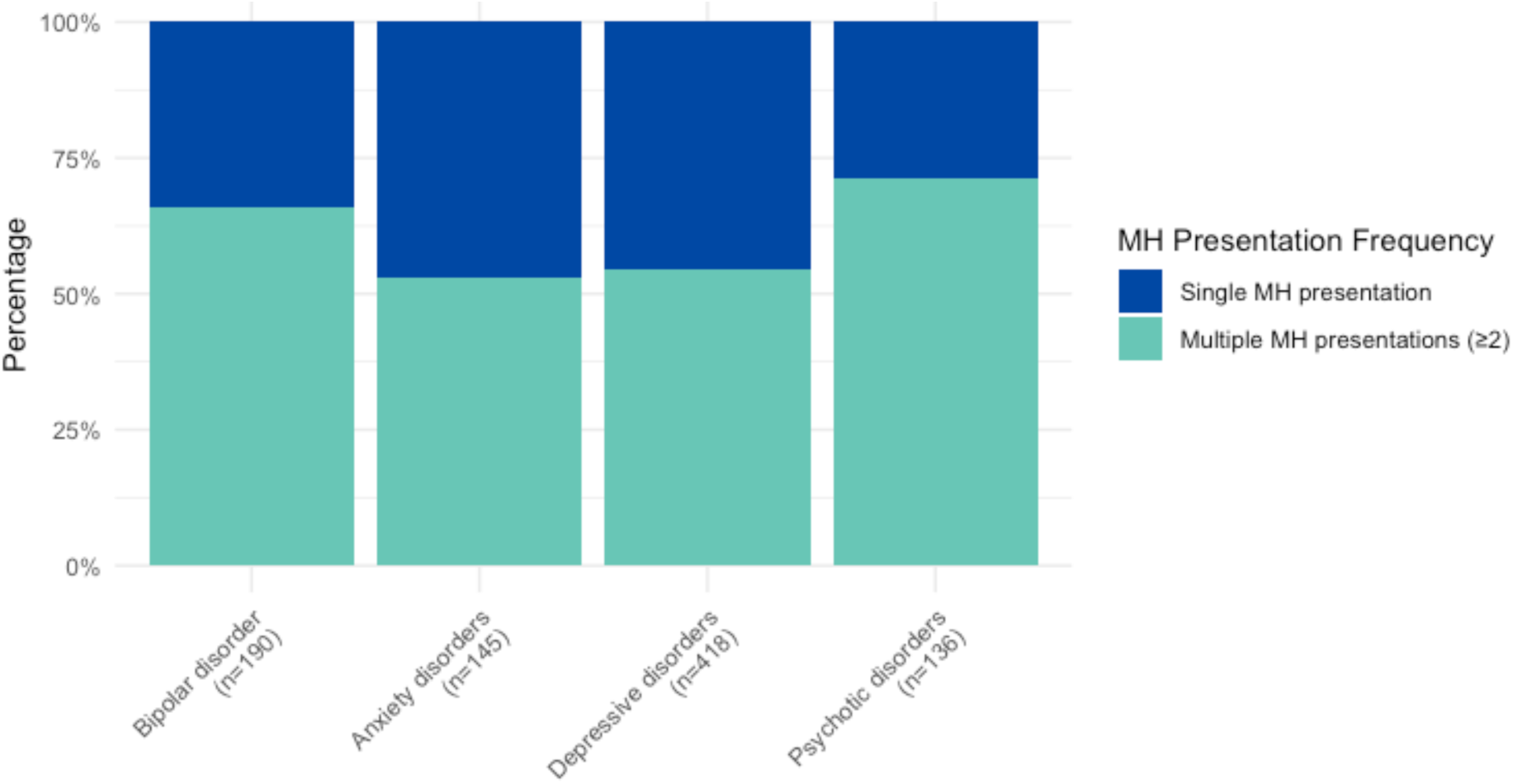
Distribution of single versus multiple mental health-related ED presentation frequency (among those with any presentation) across diagnostic groups. Note: MH = mental health-related.

**Table 5.** Odds ratios of multiple mental health-related presentations for those with bipolar disorder compared to other diagnostic groups.

| Diagnostic group | Unadjusted OR [95% CI] | Adjusted OR [95% CI]<sup>a</sup> |
| --- | --- | --- |
| Anxiety disorder | 1.71 [1.10, 2.68]* | 1.26 [0.77, 2.06] |
| Depressive disorder | 1.62 [1.13, 2.33]* | 1.36 [0.92, 2.02] |
| Psychotic disorder | 0.77 [0.47, 1.25] | 1.01 [0.59, 1.71] |

<sup>a</sup>Adjusted for age at first ED presentation, sex, clinical stage, and functioning.
Abbreviations: OR = Odds Ratio; CI = Confidence Interval.
n=875 (n=14 excluded due to 'uncertain' or missing clinical stage).
Reference group: Bipolar disorder.
\*p<.05, \*\*p<.001

However, after adjustment, these diagnostic differences were no longer significant (anxiety: OR=1.26, 95% CI [0.77, 2.06], p=.356); depressive: OR=1.36, 95% CI [0.92, 2.02], p=.130; psychotic: OR=1.01, 95% CI [0.59, 1.71], p=.975). Clinical stage and functioning were independently associated with multiple mental health-related presentations (Table S6), with more advanced clinical stage (2+ versus 1a; OR=2.39, 95% CI [1.28, 4.53], p=.007) and greater functional impairment (OR=0.96, 95% CI [0.93, 0.99], p=.013) associated with higher odds.

## 4. DISCUSSION

Building on prior evience of high ED utilisation among youth engaged in mental health services [16], our findings show that young people with BD exhibit distinct patterns of ED use compared to those with common anxiety and depressive disorders, while closely resembling those with psychotic disorders. BD is associated with greater ED use than anxiety and depressive disorders; however, several of these differences are attenuated after adjusting for clinical stage and functioning, indicating that markers of illness progression partly explain the observed differences.

Our findings emphasise that young people with BD represent a high-needs group with frequent and recurrent ED use. Almost nine in ten (87.29%) presented to an ED at least once during the 10-year study period, and they were more likely to have multiple presentations (2+) than a single presentation. ED use patterns differed between BD and anxiety and depressive disorders, with higher rates largely driven by mental health-related presentations. In contrast, individuals with BD and psychotic disorders show similar patterns across likelihood of presentation, recurrence, overall rates, and mental health-related presentations, suggesting comparably high service needs in both groups.

These findings align with previous evidence linking increasing ED use to greater mental illness severity [23] and extend prior work by demonstrating higher ED use in BD relative to anxiety and depressive disorders. This occurs despite previous reports of limited clinical differentiation between early presentations of BD versus unipolar depressive disorders in youth [24]. However, adjusted models indicate that markers of illness severity or progression (clinical stage and socio-occupational functioning) strongly predict recurrent mental health presentations to ED, attenuating diagnostic differences. This pattern suggests that recurrent ED use reflects illness progression more than diagnosis alone.

The need for repeated mental health-related ED presentations highlight gaps in community-based youth services, particularly for more severe or complex cases. Availability of community-based services does not necessarily reduce psychiatric ED use among individuals with severe mental disorders [25]. The headspace model primarily services young people with mild-to-moderate needs through brief or single-session interventions [26], yet many young people present with advanced clinical stage [27] and significant functional impairment [28]. Approximately one in five young people presenting to headspace fall into a “high complexity” subgroup [29], and emerging evidence suggests reduced effectiveness of these services for such individuals [28, 30, 31]. Young people with greater complexity may be ‘too unwell’ or disengaged for brief intervention models [32]. This contributes to the “missing middle service gap”, affecting an estimated 40% of young people who are too complex or severe for primary care [33] yet cannot access specialised services. Those who repeatedly present to EDs likely require sustained, coordinated care, which EDs cannot provide, particularly given limited access to mental health professionals in these settings [34]. Although EDs remain an essential service in situations of high acuity or concerns of safety, they are not a suitable substitute for ongoing care, highlighting the need for accessible, community-based services that have the capacity to provide care to those across the spectrum of mental ill-health.

ED presentations among those with BD are higher relative to anxiety and depressive disorders, and adjustment for illness progression does not fully explain these differences. This suggests that, while illness severity contributes to acute service use, features intrinsic to bipolar phenomenology may also drive presentations. One possibility is that BD involves recurrent episodes of mood mood instability and intensity [35], which may drive acute presentations beyond what is captured by static measures of functioning or clinical stage. Depressive episode severity is strongly influenced by mixed features and psychotic symptoms rather than by diagnostic category (i.e., unipolar versus bipolar) alone, indicating that transdiagnostic symptom dimensions are key determinants of clinical intensity [36]. Mixed features are associated with higher health service use during depressive episodes, and greater suicidality and comorbidity during manic episodes, increasing acute care needs [37]. As these features occur more frequently in BD than unipolar depressive disorders [38], they may partly explain persistent elevations in mental health-related ED presentations. Although this study did not examine specific symptom profiles, greater variability in mood episodes likely contributes to recurrent acute destabilisations and where community services cannot address rapidly escalating or complex presentations, EDs may become the default care setting. Future research should examine specific symptom profiles and mood states preceding ED presentations to inform targeted community-based supports.

An important methodological distinction, and strength, of this study is the use of longitudinal cohort data with clinically characterised diagnoses from outpatient care, rather than relying on ED-assigned diagnostic codes. Previous studies have classified depressive and bipolar presentations based on the reason for presentation to an ED [39] – that is, the primary complaint or diagnosis assigned during the ED visit itself. This approach has significant limitations: (1) it fails to capture complexity in healthcare needs which may include ED presentations for depressive episodes, alcohol of substance misuse, suicidal behaviours, or physical health issues that may be related to their underlying disorder; and (2) single-episode coding in the ED is unlikely to provide reliable diagnostic information, particularly as many BD diagnoses received in the ED are revised following further assessment [40]. By examining ED use patterns among young people included in a longitudinal cohort with well-characterised clinical information, this study provides a more comprehensive picture of the emergency service burden associated with different mental health conditions among youth.

Several limitations should be considered. First, diagnostic classification was the cross-sectional, based on the most recent service presentation and clinical notes, and does not capture the dynamic nature of mental health diagnoses in youth, where diagnostic and severity transitions occur over time [41]. Future work is needed to understand how ED presentation patterns are influenced by transitions in diagnoses and other markers of illness complexity (e.g., functional impairment) over time. Second, we could not determine the temporal sequence between ED use, diagnosis, and service engagement, limiting interpretation of care pathways. However, the focus on the relationship between established diagnoses and ED use patterns provides valuable insights into the general service needs of youth with different mental health conditions. Third, based on the available data, there are a number of confounders that we did not consider, such as BD subtypes, sub- or full-threshold disorders, co-morbid mental health diagnoses, personality (e.g. neuroticism), socio-economic status, homelessness, or medication use. Prior work shows higher ED use among those with BD type I versus type II and ‘not otherwise specified’ [42] and those receiving inadequate treatment [43], and this should be explored in future analyses. Fourth, although presentation reasons were systematically coded with high interrater reliability, they relied on administrative records and may reflect variability in documentation across EDs and/or clinicians. Fifth, the BPRR register includes data from young people attending two services across metropolitan Sydney, limiting generalisablity to regional and rural areas, where ED use is higher (particularly in young people) [44]. Sixth, this study did not assess service experiences (e.g., accessibility, satisfaction, responsiveness), which may influence care pathways. Finally, we used a complete case analysis approach for unadjusted and adjusted models, excluding participants with missing data on covariates, resulting in an analytic sample of 1778 participants who had presented to an ED. This may have introduced selection bias if missingness was associated with both diagnostic group and presentation patterns. However, sensitivity analyses re-running unadjusted models with the full sample (n=1800) showed that results did not differ meaningfully from those using the complete case sample (see Tables S7–10), indicating minimal impact of missing data.

In summary, in this youth mental health cohort, BD is associated with elevated ED use, particularly for mental health-related presentations. Young people with BD are more likely than those with anxiety or depressive disorders to present for mental health-related reasons and show patterns comparable to those with psychotic disorders. Recurrent mental health presentations are largely explained by markers of illness progression, including clinical stage and functioning, raising questions about the suitability of community-based services for more complex cases regardless of diagnosis. Taken together, these findings highlight the substantial needs of young people with BD and the importance of strengthening continuity of care and expanding community-based services to reduce reliance on emergency care.

## Author contributions

AT: Conceptualisation, data curation, investigation, methodology, project administration, formal analysis, visualisation, writing - original draft; IBH: Conceptualisation, methodology, writing – review & editing; MV: Methodology, review – reviewing & editing; NH: Methodology, writing – review & editing; CM: Project administration, writing – review & editing; JJC: Methodology, writing – review & editing; JSC: Writing – review & editing; AN: Project administration, writing – review & editing; NZ: Project administration, writing – review & editing; YJCS: Project administration, writing – review & editing; EMS: Writing – review & editing; FI: Conceptualisation, methodology, supervision, writing – review & editing.

## Data Availability

The de-identified data we analysed are not publicly available, but requests to the corresponding author for the data will be considered on a case-by-case basis.

## Acknowledgements

The authors would like to thank all the young people who participated in this study and all the staff in the Youth Mental Health team at the Brain and Mind Centre (University of Sydney), past and present, who contributed to this work.

