## Supplementary material for "Patterns of emergency department use among young people with bipolar disorder: A data linkage cohort study"

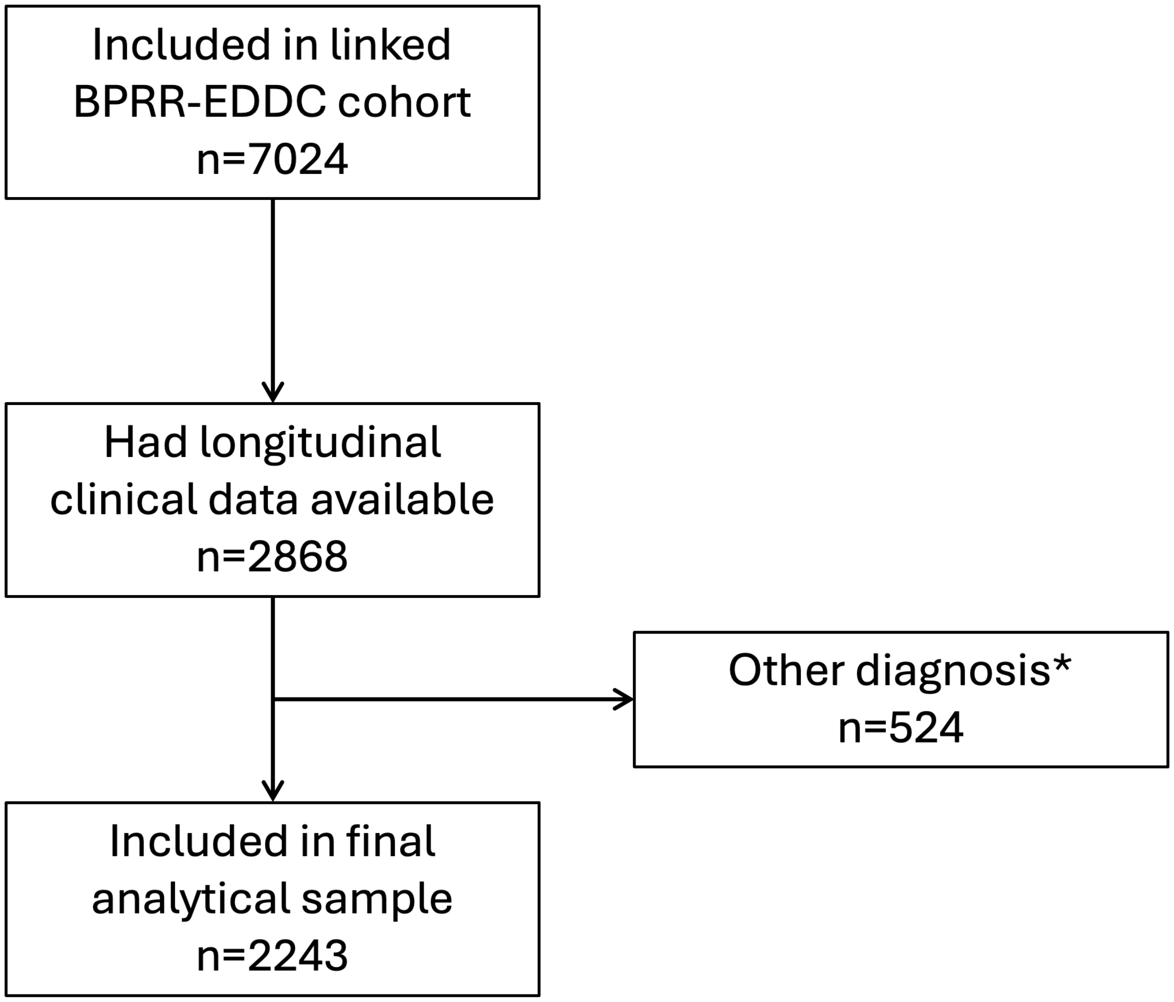

*Includes obsessive-compulsive and related disorders; trauma- and stressor-related disorders; neurodevelopmental disorder; eating disorder; personality disorder; disruptive, impulse-control and conduct disorders; substance-related and addictive disorders; uncertain diagnosis; no diagnosis.

**Supplementary Figure S1.** Flow diagram of inclusion and exclusion

**Supplementary Table S1. Demographic and clinical characteristics of those who presented to an emergency department at least once**

| Characteristic | Total sample (n=1800) | Bipolar disorder (n=309) | Anxiety disorder (n=433) | Depressive disorder (n=862) | Psychotic disorder (n=196) |
|---|---|---|---|---|---|
| Age at first ED presentation, years | 20.28 ± 4.91 | 21.38 ± 4.69 | 19.46 ± 4.70 | 19.56 ± 4.66 | 23.55 ± 5.14 |
| Sex, % female | 1117.00 (62.06) | 223.00 (72.17) | 272.00 (62.82) | 562.00 (65.20) | 60.00 (30.61) |
| Clinical stage, % yes^ | | | | | |
| Stage 1a | 249 (14.00) | 4 (1.33) | 117 (27.08) | 128 (14.95) | 0 (0.00) |
| Stage 1b | 1106 (62.20) | 142 (47.18) | 298 (68.98) | 623 (72.78) | 43 (22.75) |
| Stage 2+ | 423 (23.79) | 155 (51.50) | 17 (3.94) | 105 (12.27) | 146 (77.25) |
| SOFAS, score /100 | 62.66 ± 10.78 | 60.91 ± 12.22 | 65.08 ± 9.66 | 63.77 ± 9.86 | 55.15 ± 10.97 |

Data are presented as mean ± standard deviation for continuous variables and n (%) for categorical variables.
Abbreviations: ED = emergency department; SOFAS = Social and Occupational Functioning Scale.
^n=22 excluded due to 'uncertain' or missing clinical stage.

**Supplementary Table S2. Complete negative binomial regression results for emergency department presentation rates (adjusted for covariates)**

| Variable | IRR | 95% CI | p-value |
|---|---|---|---|
| SOFAS score | 0.97 | 0.97 – 0.98 | **<.001**** |
| *Primary diagnosis* | | | |
| Bipolar disorder (ref) | 1.00 | | |
| Anxiety disorder | 1.39 | 1.16 – 1.66 | **<.001**** |
| Depressive disorder | 1.11 | 0.95 – 1.31 | .188 |
| Psychotic disorder | 1.06 | 0.86 – 1.32 | .581 |
| *Sex* | | | |
| Female (ref) | 1.00 | | |
| Male | 1.22 | 1.09 – 1.36 | **<.001**** |
| Other | 1.43 | 0.79 – 2.58 | .242 |
| *Clinical stage* | | | |
| Stage 1a (ref) | 1.00 | | |
| Stage 1b | 0.96 | 0.82 – 1.12 | .632 |
| Stage 2+ | 0.77 | 0.63 – 0.95 | **.016*** |

Abbreviations: IRR = Incidence Rate Ratio; CI = Confidence Interval; ED = emergency department; SOFAS = Social and Occupational Functioning Scale.
n=2218 (n=25 excluded due to 'uncertain' or missing clinical stage).
Reference groups: Bipolar disorder (diagnosis), Female (sex), Stage 1a (clinical stage).
*p<.05, **p<.001

**Supplementary Table S3. Presentation reasons by diagnostic group**

| Characteristic | Total sample (n=1800) | Bipolar disorder (n=309) | Anxiety disorder (n=433) | Depressive disorder (n=862) | Psychotic disorder (n=196) |
|---|---|---|---|---|---|
| Any mental health-related presentation | 889 (49.39) | 190 (61.49) | 145 (33.49) | 418 (48.49) | 136 (69.39) |
| Mental illness | 658 (36.56) | 150 (48.54) | 107 (24.71) | 278 (32.25) | 123 (62.76) |
| Alcohol and substance misuse | 300 (16.67) | 72 (23.30) | 44 (10.16) | 147 (17.05) | 37 (18.88) |
| Suicidal behaviours and self-harm | 398 (22.11) | 77 (24.92) | 63 (14.55) | 214 (24.83) | 44 (22.45) |
| Any non-mental health presentation | 1676 (93.11) | 288 (93.20) | 405 (93.53) | 807 (93.62) | 176 (89.80) |
| Accident and injury | 992 (55.11) | 172 (55.66) | 218 (50.35) | 502 (58.24) | 100 (51.02) |
| Physical illness | 1424 (79.11) | 260 (84.14) | 338 (78.06) | 675 (78.31) | 151 (77.04) |

Data are presented as n (%)

**Supplementary Table S4. Complete logistic regression results for mental health-related presentations (adjusted for covariates)**

| Variable | OR | 95% CI | p-value |
|---|---|---|---|
| Age at first ED presentation | 0.90 | 0.88 – 0.92 | **<.001**** |
| SOFAS score | 0.96 | 0.95 – 0.97 | **<.001**** |
| *Primary diagnosis* | | | |
| Bipolar disorder (ref) | 1.00 | | |
| Anxiety disorder | 2.33 | 1.61 – 1.89 | **<.001**** |
| Depressive disorder | 1.39 | 1.02 – 1.89 | **.038** |
| Psychotic disorder | 0.87 | 0.57 – 1.33 | .523 |
| *Sex* | | | |
| Female (ref) | 1.00 | | |
| Male | 0.99 | 0.80 – 1.23 | .950 |
| Other | 1.06 | 0.35 – 3.44 | .913 |
| *Clinical stage* | | | |
| Stage 1a (ref) | 1.00 | | |
| Stage 1b | 1.84 | 1.33 – 2.56 | **<.001**** |
| Stage 2+ | 3.85 | 2.51 – 5.94 | **<.001**** |

Abbreviations: OR = Odds Ratio; CI = Confidence Interval; ED = emergency department; SOFAS = Social and Occupational Functioning Scale.

n=1778 (n=22 excluded due to 'uncertain' or missing clinical stage).

Reference groups: Bipolar disorder (diagnosis), Female (sex), Stage 1a (clinical stage).

*p<.05, **p<.001

**Supplementary Table S5. Complete logistic regression results for non-mental health presentations (adjusted for covariates)**

| Variable | OR | 95% CI | p-value |
|---|---|---|---|
| Age at first ED presentation | 0.96 | 0.92 – 0.99 | **.009*** |
| SOFAS score | 1.00 | 0.98 – 1.02 | .978 |
| *Primary diagnosis* | | | |
| Bipolar disorder (ref) | 1.00 | | |
| Anxiety disorder | 1.11 | 0.62 – 2.04 | .725 |
| Depressive disorder | 0.97 | 0.57 – 1.67 | .906 |
| Psychotic disorder | 1.41 | 0.73 – 2.70 | .307 |
| *Sex* | | | |
| Female (ref) | 1.00 | | |
| Male | 0.72 | 0.50 – 1.04 | .075 |
| Other | 0.96 | 0.18 – 17.67 | .969 |
| *Clinical stage* | | | |
| Stage 1a (ref) | 1.00 | | |
| Stage 1b | 0.68 | 0.36 – 1.21 | .210 |
| Stage 2+ | 0.84 | 0.38 – 1.79 | .657 |

Abbreviations: OR = Odds Ratio; CI = Confidence Interval; ED = emergency department; SOFAS = Social and Occupational Functioning Scale.
n=1778 (n=22 excluded due to ‘uncertain’ or missing clinical stage).
Reference groups: Bipolar disorder (diagnosis), Female (sex), Stage 1a (clinical stage).
*p<.05, **p<.001

**Supplementary Table S6. Complete logistic regression results for single versus multiple mental health-related presentations (adjusted for covariates)**

| Variable | OR | 95% CI | p-value |
|---|---|---|---|
| Age at first ED presentation | 0.96 | 0.93 – 0.99 | **.013*** |
| SOFAS score | 0.97 | 0.95 – 0.98 | **<.001**** |
| *Primary diagnosis* | | | |
| Bipolar disorder (ref) | 1.00 | | |
| Anxiety disorder | 1.26 | 0.77 – 2.06 | .356 |
| Depressive disorder | 1.36 | 0.92 – 2.02 | .130 |
| Psychotic disorder | 1.01 | 0.59 – 1.71 | .975 |
| *Sex* | | | |
| Female (ref) | 1.00 | | |
| Male | 1.08 | 0.80 – 1.47 | .614 |
| Other | 1.33 | 0.31 – 6.67 | .707 |
| *Clinical stage* | | | |
| Stage 1a (ref) | 1.00 | | |
| Stage 1b | 1.28 | 0.75 – 2.19 | .367 |
| Stage 2+ | 2.39 | 1.28 – 4.53 | **.007*** |

Abbreviations: OR = Odds Ratio; CI = Confidence Interval; ED = emergency department; SOFAS = Social and Occupational Functioning Scale.
n=875 (n=14 excluded due to ‘uncertain’ or missing clinical stage).
Reference groups: Bipolar disorder (diagnosis), Female (sex), Stage 1a (clinical stage).
*p<.05, **p<.001

**Supplementary Table S7. Unadjusted negative binomial regression results on complete sample (n=2243)**

| Diagnostic group | IRR | 95% CI | p-value |
|---|---|---|---|
| Anxiety disorder | 1.85 | 1.56 – 2.18 | **<.001**** |
| Depressive disorder | 1.34 | 1.15 – 1.55 | **<.001**** |
| Psychotic disorder | 0.90 | 0.73 – 1.11 | .317 |

Abbreviations: IRR = Incidence Rate Ratio; CI = Confidence Interval.
*p<.05, **p<.001

**Supplementary Table S8. Unadjusted odds ratios of mental health-related presentations on complete presented sample (n=1800)**

| Presentation reason | Diagnostic group | OR | 95% CI | p-value |
|---|---|---|---|---|
| Any mental health-related presentation | Anxiety disorder | 3.17 | 2.34 – 4.31 | **<.001**** |
| | Depressive disorder | 1.70 | 1.30 – 2.21 | **<.001**** |
| | Psychotic disorder | 0.70 | 0.48 – 1.03 | .071 |
| Any non-mental health presentation | Anxiety disorder | 0.95 | 0.53 – 1.72 | .859 |
| | Depressive disorder | 0.93 | 0.56 – 1.61 | .799 |
| | Psychotic disorder | 1.56 | 0.82 – 2.97 | .175 |

Abbreviations: OR = Odds Ratio; CI = Confidence Interval.
*p<.05, **p<.001

**Supplementary Table S9. Unadjusted odds ratios of single vs multiple mental health-related presentations on complete sample (n=889)**

| Diagnostic group | OR | 95% CI | p-value |
|---|---|---|---|
| Anxiety disorder | 1.70 | 1.09 – 2.65 | **.019*** |
| Depressive disorder | 1.60 | 1.13 – 2.30 | **.009*** |
| Psychotic disorder | 0.77 | 0.48 – 1.24 | .291 |

Abbreviations: OR = Odds Ratio; CI = Confidence Interval.
*p<.05, **p<.001

**Supplementary Table S10. Unadjusted odds ratios of individual mental health-related presentations on complete presented sample (n=1800)**

| Presentation reason | Diagnostic group | OR | 95% CI | p-value |
|---|---|---|---|---|
| Alcohol and substance misuse | Anxiety disorder | 2.69 | 1.79 – 4.06 | **<.001**** |
| | Depressive disorder | 1.48 | 1.07 – 2.02 | **.016*** |
| | Psychotic disorder | 1.31 | 0.84 – 2.05 | .240 |
| Mental illness | Anxiety disorder | 2.87 | 2.11 – 3.94 | **<.001**** |
| | Depressive disorder | 1.98 | 1.52 – 2.58 | **<.001**** |
| | Psychotic disorder | 0.56 | 0.39 – 0.81 | **.002*** |
| Suicidal behaviours and self-harm | Anxiety disorder | 1.95 | 1.35 – 2.83 | **<.001**** |
| | Depressive disorder | 1.00 | 0.74 – 1.35 | .974 |
| | Psychotic disorder | 1.15 | 0.75 – 1.76 | .526 |

Abbreviations: OR = Odds Ratio; CI = Confidence Interval.
*p<.05, **p<.001
